# Effects of 2025-2026 U.S. health and science policy changes on academic pulmonary and critical care medicine: an ASPIRE-PCCM prospective survey study

**DOI:** 10.64898/2026.09.25.26364007

**Authors:** Theodore J Iwashyna, Yvette Shu, Catherine K Ettman, Elizabeth M Viglianti, David Furfaro, Kristina Montemayor

**Affiliations:** School of Medicine, Johns Hopkins University, Baltimore, Maryland, USA; Bloomberg School of Public Health, Johns Hopkins University, Baltimore, Maryland, USA; Krieger School of Arts and Sciences, Johns Hopkins University, Baltimore, Maryland, USA; School of Medicine, University of Michigan, Ann Arbor, Michigan, USA; Beth Israel Deaconess Hospital, Boston, Massachusetts, USA

**Author notes:** **Corresponding Author:** Theodore J Iwashyna, 1830 E Monument Street, Room 519, Baltimore, Maryland, 21287. **Authorship Statement:** TJI and KM conceived of the study. TJI, CKE, EMV, DF, and KM developed the study methods. TJI and KM organized data collection. TJI and YS analyzed the data. TJI drafted the manuscript. All others contributed substantially to the interpretation, critically reviewed the manuscript, and approved of its final form. **Funding:** No external funding was received for this project. **Disclosures:** TJI is a paid consultant and potential expert witness for Roots Community Health of Oakland, California, on matters of pulse oximetry not related to the current manuscript. **IRB Review:** This proposal was deemed not human subjects research by the Johns Hopkins University on the basis that it was a survey of professionals expressing views on their professional activity, with the option to opt out of any question.

**Keywords:** health policy, science policy, vaccines, immigration, survey

## Abstract

**Background:** The rapidly evolving United States (U.S.) health and science policy landscape in 2025 may have heterogeneous effects on academic medicine, biomedical research, physician workforce development and patient care.

**Research Question:** How do academic pulmonary and critical care medicine (PCCM) physicians perceive U.S. health and science policy changes and which changes do they consider most influential on research, trainee education and clinical care?

**Study Design and Methods:** We conducted a prospective survey from March 27, 2026 through April 29, 2026 among selected faculty at U.S. academic PCCM programs as part of the Alliance of Scholars and Professionals for Innovation, Research, and Education in Pulmonary and Critical Care Medicine (ASPIRE-PCCM).

**Results:** Ninety-four of 99 ASPIRE-PCCM panel members completed the survey (response rate 95%), representing 18 of 19 ASPIRE-PCCM wave 1 institutions. All respondents reported the rate of changes in U.S. federal health and science policy in the first half of 2025 as 6 or higher on a scale of 1 (“typical pace of change”) to 10 (“most change in my lifetime”), with 89 (96%) evaluating the changes as harmful or very harmful. Respondents identified cancellation of research grants, disruptions and changes to NIH scientific review processes, and changes to visa and immigration policies as key factors affecting early career recruitment. Reported negative effects on quality of care included increased vaccine hesitancy, changing reimbursement structures, and diminished research capacity. State-level and non-governmental efforts, including professional society advocacy, institutional bridge funding and personal advocacy efforts were identified as partially mitigating harms. Despite these concerns, 72% of respondents reported that would still pursue careers in PCCM.

**Interpretation:** Academic pulmonary and critical care physicians reported challenges following recent U.S. health and science policy changes that may be mitigated, but not neutralized, by institutional, professional-society, and individual actions.

---

In the United States (U.S.) 2025 and early 2026 have been marked by substantial policy changes affecting many facets of academic medicine. The uncertainty surrounding these changes, as well as the implementation, temporary restraining orders, and downstream sequelae may impact how medical care is delivered and studied.

There is reason to believe that the impacts of policy changes may be heterogeneous across medical specialties.^1^ Pulmonary and Critical Care Medicine (PCCM) may be particularly susceptible to benefits and harms given its heavy reliance on hospital funding streams, longstanding dependence on federally funded scientific research,^2^ and care that includes a wide-range of patients with conditions sensitive to access to care,^3,4^ multi-disciplinary diagnostic^5-7^ evaluations, environmental exposures, and costly therapies. The field is also characterized by a rapid pace of bench-to-bedside advances. These challenges emerge as PCCM specialists report high rates of burnout even relative to other physicians.

Therefore, we conducted a prospective quantitative and qualitative survey of academic faculty participating in the Alliance of Scholars and Professionals for Innovation, Research, and Education in Pulmonary and Critical Care Medicine (ASPIRE-PCCM). We sought to identify the overall impacts of these policy changes on aspects of the tripartite mission of care, education, and research, and to further identify the changes they felt were most consequential.

## METHODS

### Survey Panel Recruitment

The ASPIRE-PCCM longitudinal survey panel was developed to leverage the knowledge, opinions, and self-reported practice patterns of physicians in PCCM who are assistant, associate, or full professors at U.S. academic medical centers that also serve as research and referral centers. Guided by an Advisory Committee, the panel was recruited in 3 stages. First, a list of PCCM programs with NHLBI T32 programs administered by a Lung Section Program Officer in 2025 was developed. This was de-duplicated, and pediatric programs were removed; it was then supplemented by adding other adult PCCM divisions recognized for comprehensive clinical training, scientific inquiry, and fellowship educational reputation. We intentionally selected and developed a prioritized list of programs to ensure a range of geography and populations served. Second, we attempted to recruit a site lead for programs, often a fellowship director or other faculty member with broad knowledge of the division (See **Appendix 1** for list of sites and site leads). Third, approximately 5 faculty respondents were recruited by site leads for each site. Site leads were instructed to select faculty recognized for excellence in clinical care, research, and education, who would be responsive members of the survey panel and reflected the diversity of their faculty along multiple axes including clinical area of focus, demographics, and rank.

Panel members committed to timely completion of up to four 30-minute surveys per year; each survey is referred to as a wave and this report describes components of the first wave. No financial compensation was provided. Participants received early access to survey results and the opportunity to suggest future survey modules. Panel members who fail to complete more than one survey each year are informed that they may be replaced. Sites with regular missing data will undergo remediation support and if challenges persist despite these efforts, alternative site arrangements may be considered. We intend that participants who leave their respective programs will in general be invited to remain in the panel and potentially replaced at their former institution.

### Survey Item Development and Administration

Survey modules were developed through an iterative process and will continue to evolve over subsequent waves. For each proposed module, a reporting shell was developed to guide dissemination as a distinct manuscript. Reporting shells were reviewed and revised by the ASPIRE-PCCM leads and Advisory Committee for significance, mission-concordance, portfolio balance, rigor, and feasibility. ASPIRE-PCCM Wave 1 includes three modules: (1) health and science policy changes; (2) care of critically ill oncology patients, including immune-checkpoint inhibitor pneumonitis; and (3) pulmonary procedures and learner performance. The present manuscript reports findings from the health and science policy changes module.

We developed by survey items collaboratively within the team. Items about health policy drew on published reviews of 2025 policy changes^.8-10^ A shared demographics module was developed drawing on a published instrument by Costa, Gershengorn and colleagues,^11^ among others.

Survey items were iteratively refined by feedback with survey experts, epidemiologists, and clinicians; coded and tested in Redcap survey administration software; underwent cognitive^12-14^ interviewing with at least 5 relevant informants; and finalized after review by ASPIRE-PCCM leads and staff. As a standard practice, all surveys include a free text option to allow panel members to comment further on any issues brought up by the survey items, and to allow opt-in for optional follow-up qualitative interviews about the topics of the survey.

The complete survey instrument is provided in **Appendix 2**. The study protocol and survey design underwent review by an Institutional Review Board and were deemed exempt. Informed consent was obtained at the first survey for each panel member, including careful explanations of the following confidentiality provisions. Survey responses were treated as confidential but not anonymous to allow longitudinal linkage over time and collection of demographic characteristics. Participants were assured that no individually identifiable data will be released outside of the study team without legal or governmental compulsion. Consent information was provided on the first page of the survey and documented through a required electronic consent question.

Surveys were administered through university-administered IRB-approved survey software, Redcap. Site recruitment began following IRB-approval. Individualized surveys were sent to panel members via their preferred emails, with automated reminders at 3 and 7 days later, and intermittently thereafter. Site leads are given information about which of their site members have responded to allow the site lead to support their panel members in providing responses. Site leads do not have access to survey responses, only to the fact or not of survey completion at a given timepoint. Surveys were completed from March 27, 2026 through April 29, 2026.

### Statistical and Free Text Analyses

Responses were summarized using standard categorical and continuous variables. Respondents’ region was classified into Census category by the location of the primary university campus. Fischer’s exact test was used whenever any cross-tabulation included a cell size less than zero. The number of responses to any item is reported to evaluate missingness; because missingness was rare, casewise deletion was used rather than multiple imputation.

Free text responses were analyzed inductively by YS and TJI, identifying and grouping common themes, and then presenting particularly illustrative quotes.

No large language model or so-called artificial intelligence tools were used in the production of this manuscript.

## RESULTS

We identified 45 potential sites. 25 sites were approached for potential participation; 19 agreed to participate (**Appendix 1**), were able to complete any review processes for wave 1, and provided at least one survey panel member’s contact information, for a total of 99 potential respondents at this first ASPIRE-PCCM wave. Of these, 94 respondents from 18 sites completed the survey, a 95% response rate.

Of respondents, their median age was 42; 38 were assistant professors or equivalent, 34 associate professors or equivalent, and 14 full professors; 78 considered themselves involved in research; and (not mutually exclusively) 83 involved in education. Other demographics are in **Table 1**. Responding sites were from all Census regions: 6 in the south, 5 each in the northeast and midwest, 2 in the west.

**Table 1:** Participant Demographics.

|  |  |
| --- | --- |
| Year of Birth (Median, IQR) | 1984 (1975-1986) |
| Gender (n, %) |  |
| Male | 41, 45% |
| Female | 51, 55% |
| Not Reported | 2, 2% |
| Race (Self-reported, n, %) |  |
| American Indian or Alaskan Native | 1, 1% |
| Asian | 22, 24% |
| Black or African American | 2, 2% |
| Native Hawaiian or Pacific Islander | 0, 0% |
| White | 69, 74% |
| Prefer not to answer | 2, 2% |
| Write-In | 1, 1% |
| Not Reported | 1, 1% |
| Ethnicity (Self-reported, n, %) |  |
| Hispanic | 3, 3% |
| Not Hispanic | 89, 97% |
| Not Reported | 2, 2% |
| Rank (n, %) |  |
| Instructor | 4, 4% |
| Assistant professor | 38, 42% |
| Associate professor | 34, 38% |
| Full professor | 14, 16% |
| Not Reported | 4, 4% |
| Consider themselves involved in... (n, %, not mutually exclusive) |  |
| Research | 78, 85% |
| Medical Education | 83, 90% |
| Time Spent in Clinical Care (Median, IQR) |  |
| As percentage of clinical effort | 50 (25-65) |

Overall, all respondents reported the rate of changes in US federal health and science policy in the first half of 2025 as 6 or higher on a scale of 1 (typical pace of change) to 10 (most change in my lifetime), with a majority rating it 9 (n=19 (20%)) or 10 (n=41, 44%) with 2 not responding. 77 (82%, 1 not responding) felt it was “hard” or “very hard” to stay up to date on changes that were relevant to them. No respondents rated the changes as overall “very beneficial” or “beneficial”, 2 (2%) rated the changes “on balance neutral”, and 89 (96%) rated the changes as “harmful” or “very harmful” (**Figure 1**). Despite this, 68 (72%) felt these changes would not make them less likely to choose pulmonary and critical care medicine; 1 felt it would make them more likely to choose; 1 did not respond; and 24 (26%) felt that it would make them less likely to choose PCCM.

**Figure 1.**
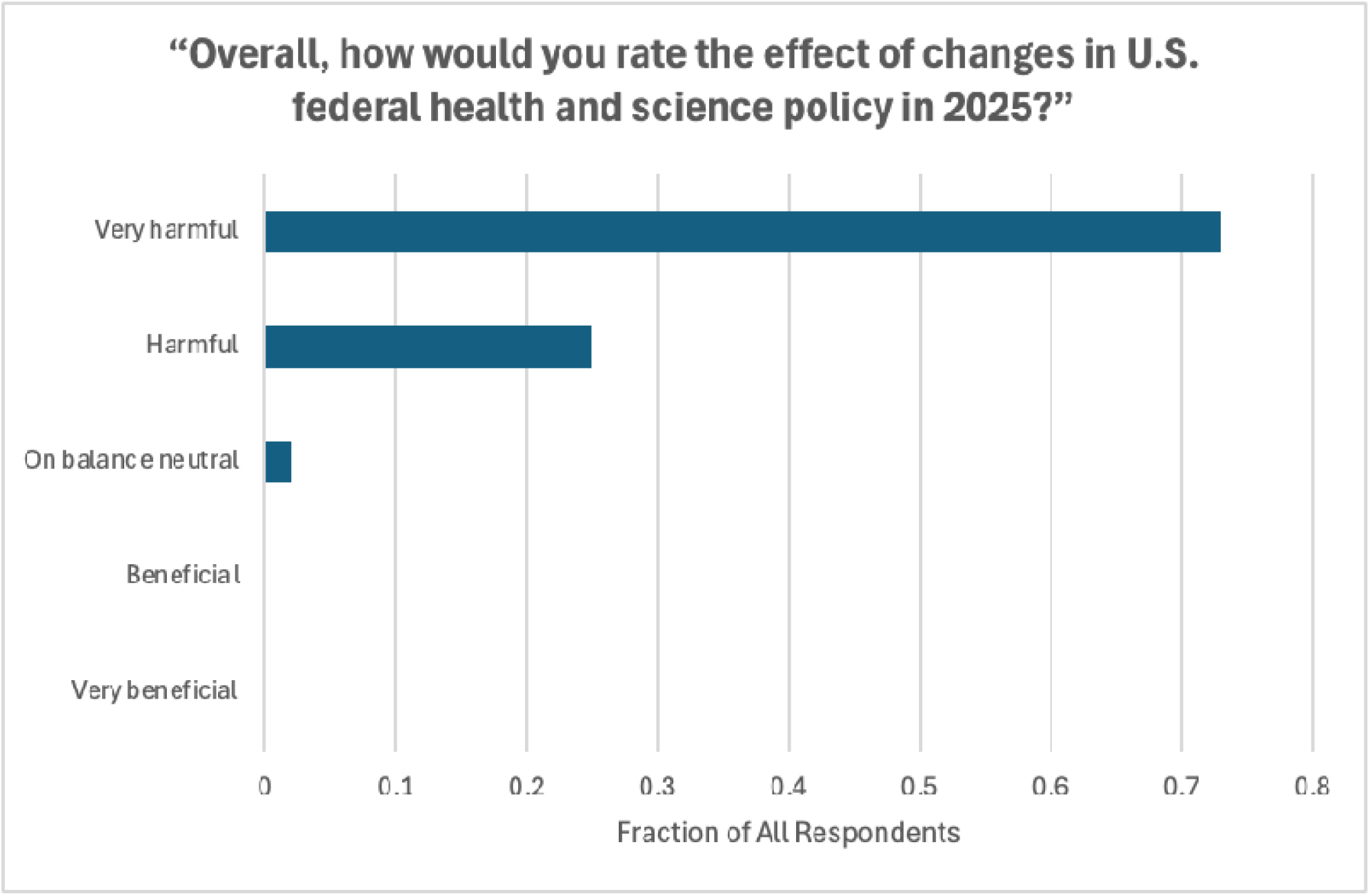
Responses to “Overall, how would you rate the effect of changes in U.S. federal health and science policy in 2025?”

### Impacts on Ability to Recruit and Retain to Fellowship and Early Faculty Positions in Pulmonary and Critical Care Medicine

Overall, most respondents (82%, 2 non-respondents) reported that changes had influenced their ability to recruit outstanding career individuals to pulmonary and critical care medicine; all rated changes as “harmful” or “very harmful”.

Respondents were provided a list of 17 policy changes identified in the literature review, (**Table 2**) and asked to select the 3 they considered most influential for early career recruitment. More than half-selected “cancellation of research grants”; “NIH scientific review disruptions and changes”; “visa and immigration policy changes”. Also commonly selected as influential, by approximately one-third, were “indirect cost formula proposed changes and uncertainty”; “Suspension of and lawsuits around DEI activities”. Most respondents (79, 85%, 2 non-respondents) felt that these policy changes were already influencing recruitment and retention. Asked specifically about the impact on success as physician-scientists, the picture was quite similar.

**Table 2:** Selection of Policy Changes as “Top 3 Most Consequential”. Sorted by most consequential for recruitment and retention. The most commonly selected answers in each column are bolded.

|  | Most<br>consequential<br>for<br>recruitment<br>and retention | Most<br>consequential<br>for your<br>ability to<br>deliver high-quality patient<br>care |
| --- | --- | --- |
| **Cancellation of research grants** | **62 (66.7%)** | 9 (9.8%) |
| **NIH scientific review disruptions and changes** | **56 (60.2%)** | 16 (17.4%) |
| **Visa and immigration policy changes** | **49 (52.7%)** | 14 (15.2%) |
| Indirect cost formula proposed changes and uncertainty | 32 (34.4%) | 10 (10.9%) |
| Suspension of and lawsuits around DEI activities | 29 (31.2%) | 7 (7.6%) |
| **Medicaid funding changes** | 7 (7.5%) | **52 (56.5%)** |
| **Vaccine policy changes** | 6 (6.5%) | **45 (48.9%)** |
| ICE deportation and enforcement activities | 6 (6.5%) | 11 (12.0%), |
| **Medicare funding changes** | 5 (5.4%) | **46 (50.0%)** |
| LGBTQ+ care changes | 5 (5.4%) | 5 (5.4%) |
| Government data availability | 4 (4.3%) | 13 (14.1%) |
| MAHA commission recommendations | 4 (4.3%) | 14 (15.2%) |
| Cancellation of domestic care delivery programs | 3 (3.2%) | 21 (22.8%) |
| Dobbs-related changes in access to reproductive care | 3 (3.2%) | 7 (7.6%) |
| EPA pollution program changes | 3 (3.2%) | 5 (5.4%) |
| Suspensions of overseas aid and related care delivery | 2 (2.2%) | 0 (0.0%) |
| Ability of government employees to publish scientific articles | 2 (2.2%) | 1 (1.1%) |

### Impact on Quality of Care

Overall, respondents felt that the impacts of policy change on their ability to provide high-quality PCCM care were more severe than the impact on recruitment and retention (p=0.001 by Fisher’s exact test), with only 8 reporting changes on “balance neutral” and none reporting the changes as positive. (**Figure 2**) 79 (85%) felt that changes had already occurred. Approximately half of respondents identified Medicaid funding changes, Medicare funding changes, and / or vaccine funding changes as most influential for their ability to provide high quality care, but answers were widely spread across all 16 policies.

**Figure 2.**
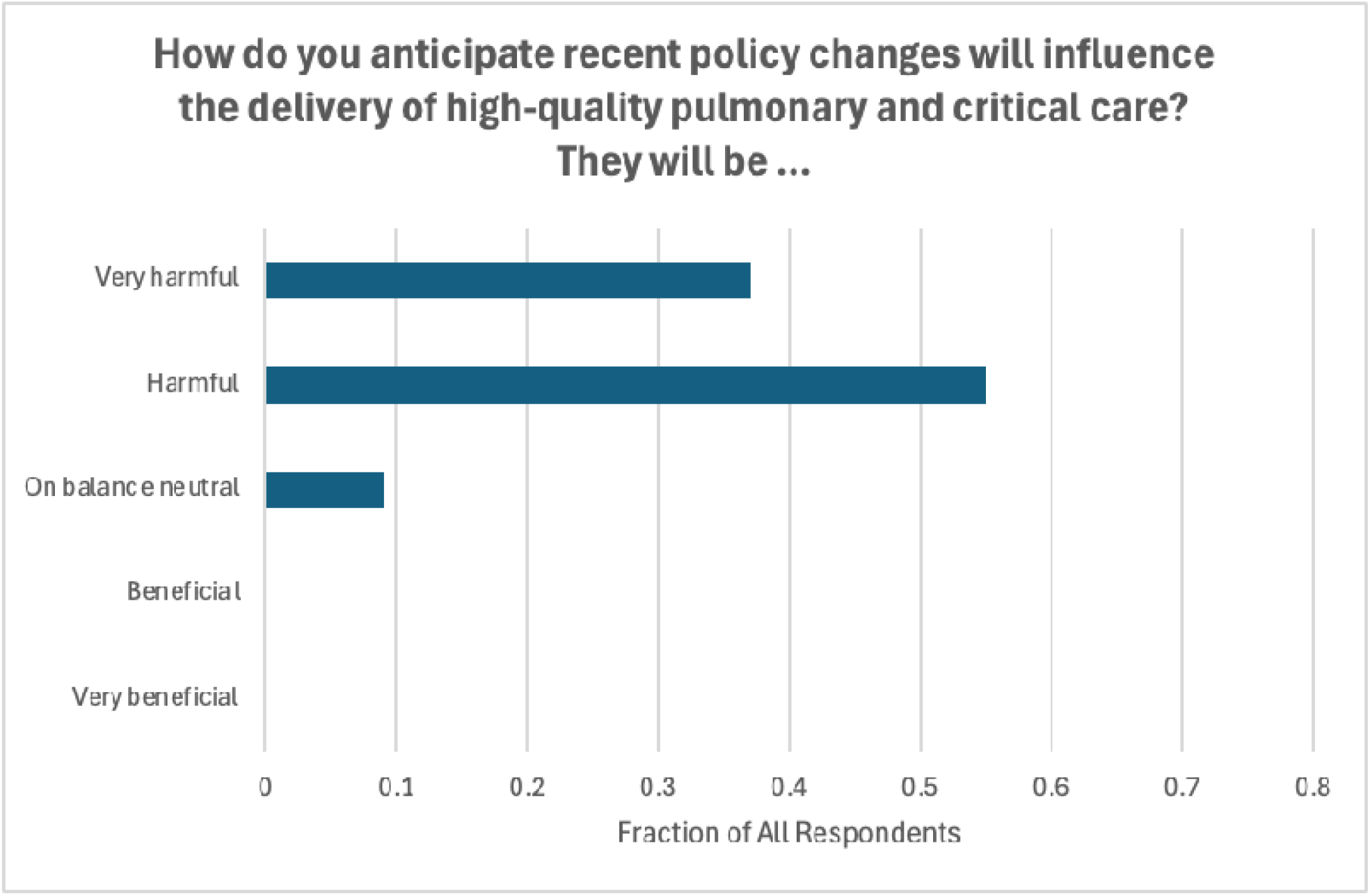
Responses to “How do you anticipate recent policy changes will influence the delivery of high-quality pulmonary and critical care? They will be…”.

More than three-quarters of respondents provided examples of adverse impacts on their ability to deliver high-quality care. Many wrote with personal poignancy to highlight concerns regarding recurring themes about:

1. vaccine hesitancy and suspicion in scientific research, which they felt were actively, and would continually, leading to outbreaks of preventable illnesses. One respondent wrote: “Patients are increasingly hesitant about vaccines, and the increased burden of shared decision-making around vaccines takes away from time spent implementing evidence-based best practices.”
2. reduced Medicaid/Medicare/insurance coverage that, they felt, will decrease hospital funding and patient accessibility. As one wrote, “More uninsured/underinsured patients if Medicaid eligibility or funding changes resulting in delayed presentations, more severe disease at ICU admission.”
3. changing immigration policies that, they felt, were detrimental to patients and staff. One respondent wrote: “A concrete example is that a patient in ICE custody was recently brought in for outpatient care and did not feel comfortable discussing their HIV status and anything related to sensitive medical topics as the agents refused to leave the room.”
4. decreased government funding in research will, they felt, further marginalize populations and leave hospitals unprepared for future issues, such as climate change impacts onto patient health. One wrote: “my institution has already had to let some young scientists go and pipeline for replacing them will be difficult to fill if/when circumstances change brain drain to other countries has occurred teaching/research time has been cut in half for CEs (20% --> 10%) across the board, affecting recruitment, morale, and research/advocacy activities.” Another wrote of the “failure to mitigate an already-increasing set of climate change related negative health outcomes (e.g., wildfire and heat related cardiorespiratory pathologies)”

### Satisfaction with Response from Professional Societies

Respondents reported membership in a median of 1-2 of the 5 queried professional organizations. (Participation in the American College of Physicians was low and so it was excluded from further analyses.) The most common evaluation of the professional societies responses to policy changes was that the respondents were unaware of their activity. Evaluations of the professional societies as “too confrontational” were rare. (**Table 3**)

**Table 3:** Evaluation of Policy Approaches of Select Medical Professional Organizations. ACCP = American College of Chest Physicians; ATS = American Thoracic Society; SCCM = Society of Critical Care Medicine; AMA = American Medical Association.

|  | Unaware of<br>Activities | Too<br>Confrontational | About Right | Too Agreement-Oriented | No Response |
| --- | --- | --- | --- | --- | --- |
| ACCP | 47 (54%) | 0 (0%) | 23 (26%) | 17 (20%) | 7 (7%) |
| ATS | 28 (31%) | 2 (2%) | 31 (34%) | 30 (33%) | 3 (3%) |
| SCCM | 59 (73%) | 0 (0%) | 8 (10%) | 14 (17%) | 13 (14%) |
| AMA | 50 (61%) | 1 (1%) | 9 (11%) | 22 (27%) | 12 (13%) |

### Harm Mitigation Strategies

Respondents were asked an open-ended question about strategies they knew of or had adopted to “maximize the positive impact or minimize the negative impacts of health and science policy changes in 2025”. Free-text responses emphasized harm mitigation. Some respondents noted that states and non-governmental organizations have issued counter-interventions to federal policies, especially around vaccines—as one wrote, “The American Academy of Pediatrics (AAP), took a stand on vaccine policy and broke with the current administration to publish their own (evidence-based) childhood vaccine schedule” and another praised the “West Coast Health Alliance”. There were mentions of increased political engagement, with one saying “I have participated in in person and phone lobbying of [state-redacted to preserve anonymity] politicians.”

Others noted local institutional responses, including bridge funding and clarification of policy implementation; as one respondent wrote “Hospital administration at our safety net hospital have done a good job sending out emails preparing trainees on what to say if ICE does come to talk to staff regarding patient data.” Finally, a number emphasized personalistic approaches and recommitment to themselves and their mission, writing that they “continue the work that I am most passionate about and encourage others to do the same”; “help trainees focus on the process. Refocus on core values and goals. Diversify funding streams as best able”; and “keep calm and carry on.”

## DISCUSSION

Physician PCCM faculty committed to excellence in clinical care, scientific research, and education, at academic medical centers, perceive widespread, rapid, consequential negative effects of health and science policy changes in the United States in 2025-2026. The instability and modification of funding commitments, and rapid changes in DEI and visa policies have been seen as already reducing the quality of science being produced and the ability to educate physician-scientists. Changes in vaccine policy, promotion of medical disinformation, and intrusive immigration enforcement are perceived as already reducing the ability to deliver the highest quality of pulmonary and critical care.

There has been a growing body of work about the impacts of policy and science changes on all aspects of the long arc of scientific research, including in the previous administrations.^15^ Impacts on clinical trials have been documented, with perhaps 1 in 30 clinical trials disrupted.^16^ A recently published analysis of disruptions in clinical trials and other federal scientific grants noted that “statistical patterns show that early-career investigators—assistant professors, postdoctoral scholars, trainees, and graduate students—were disproportionately affected, as were women.”^17^ Perceptions of substantial adverse impact by K-awardees have been described.^18,19^ The present data suggest impacts in pulmonary and critical care are perceived to be at least as significant as these general bioscience impacts, and with wide-ranging concerns about care quality—not limited to those previously noted from abortion bans^20^—not only in the future but in the present.^21^

Evaluations of professional society responses to changes have been more limited, albeit building on recent work.^22,23^Some but not all professional societies relevant to pulmonary and critical care medicine made strong public statements in support of freedom of the press on matters of academic and scientific publishing.^24,25^ There has been more consensus on matters of vaccine safety and promotion.^26-30^ The current survey suggests that, at least among ASPIRE-PCCM’s selected sample of physician faculty, there is little concern that professional societies have been too vocal, and some appetite for a more aggressive stand.

These results have distinctive strengths and weaknesses that need to be considered in their interpretation. There is no universally accepted definition of academic pulmonary and critical care medicine; therefore, we used a site-based recruiting framework based on but not exclusively limited to NHLBI Lung Section T32 receipt. Within academic pulmonary and critical care programs, there is heterogeneous engagement among faculty in the tripartite mission; faculty exclusively focused on excellence in one area have been described. Therefore, we recruited site leads with day-to-day knowledge of potential respondents, and asked them to “select faculty recognized for excellence in clinical care, research, and education.” Respondents were also committing to longitudinal participation in the ASPIRE-PCCM program. Thus the survey is neither intended to be, nor should be, the same as other potential sampling frames—such as all faculty member, all university-hospital affiliated clinicians, professional society membership, or community-practicing clinicians. The limited sample size is intentionally balanced by the high response rate, and we plan that in future ASPIRE-PCCM waves, additional sites will be recruited.

Likewise, this survey reflects the views of its expert respondents, and is not a formal policy evaluation with quantitative counterfactuals, nor were the impacts of policy changes randomly rolled in, as in a stepped-wedge cluster randomization design. We report only what these respondents perceive as happening and their causal attributions; we can here neither prove nor disprove them.

In conclusion, these data reveal substantial concerns among academic pulmonary and critical care physician faculty that recent changes in U.S. federal health and science policy have already negatively impacted the health and care of Americans, and will have increasingly negative impacts on future health care and scientific productivity unless there are changes.

## Supporting information

Appendix 1 List of Participating Sites

## Data Availability

All data produced in the present study are available upon reasonable request to the authors

## Acknowledgments

The authors wish to thank Taylor Bernstein for her expert project management, and the survey participants for their time. We thank Dr. Josh Fessel for his expert input into the development of the sampling frame.

