## Appendix 1 List of Participating Sites for "Effects of 2025-2026 U.S. health and science policy changes on academic pulmonary and critical care medicine: an ASPIRE-PCCM prospective survey study"

**Appendix 1**. Site Leads & Institution

| **Institution** | **Name** |
| --- | --- |
| Indiana University | Gabe Bosslet |
| Johns Hopkins University | Ashraf Fawzy |
| Mass General Brigham | Kathryn Hibbert |
| New York University | Deepak Pradhan |
| Northwestern University | Catherine A. Gao |
| Oregon Health & Science University | Katrina Hauschildt & Kelly Vranas |
| Stanford University | Andrea Jonas |
| University of Colorado | Tom Valley |
| University of Kentucky | Ashley Montgomery-Yates |
| University of Miami | Chris Mallow |
| University of Michigan | Jake Mcsparron |
| University of Minnesota | Katie Pendleton |
| University of Pennsylvania | C Jessica Dine |
| University of Virginia | Kyle Enfield |
| University of Wisconsin | Jacky Kruser |
| Vanderbilt University | Carla Sevin |
| Wake Forest University | Abigail Koch |
| Yale University | Lauren Ferrante |
